# *CarpeDiem*, a per-day clinical parameters and pneumonia adjudication dataset for critically ill patients with suspected pneumonia

**DOI:** 10.64898/2026.03.10.26348076

**Authors:** Catherine A. Gao, Nikolay S. Markov, Mengjia Kang, Luke V. Rasmussen, Wan-Ting Liao, Anna Pawlowski, Prasanth Nannapaneni, Vijeeth Guggilla, Helen K. Donnelly, Rebecca K. Clepp, Chiagozie Pickens, Nandita Nadig, Thomas Stoeger, Dan Schneider, Justin Starren, Theresa Walunas, G.R. Scott Budinger, Richard G. Wunderink, Alexander V. Misharin, Benjamin D. Singer, The NU SCRIPT Study Investigators

**Affiliations:** Division of Pulmonary and Critical Care Medicine, Department of Medicine, Northwestern University Feinberg School of Medicine, Chicago, IL, USA; Division of Biostatistics and Informatics, Northwestern University Feinberg School of Medicine, Chicago, IL, USA; Northwestern Medicine Enterprise Data Warehouse; Division of General Internal Medicine, Department of Medicine, Northwestern University Feinberg School of Medicine, Chicago, IL, USA

**Author notes:** GRSB, RGW, AVM, and BDS are co–senior authors.

## Abstract

Many analyses of critically ill patients focus on admission features and discharge outcomes, overlooking daily events and intercurrent complications. Pneumonia is a syndromic entity that is difficult to adjudicate outcomes without clinical expertise. Here, we examine clinical features on a day-by-day basis, mirroring the practice of daily multidisciplinary rounds, with annotated pneumonia episodes and outcomes. The Successful Clinical Response in Pneumonia Therapy (SCRIPT) *CarpeDiem* Dataset includes 21,931 patient-hospital-days from 704 patients enrolled in the SCRIPT study between June 2018 and May 2023. These patients were all receiving mechanical ventilation and underwent bronchoalveolar lavage to diagnose and establish the etiology of their pneumonia or an alternative diagnosis. The dataset captures patient demographics, daily clinical parameters, including vital signs, laboratory results, and mechanical support data, detailed pneumonia episode results adjudicated by critical care physicians, and outcomes. All data are de-identified per HIPAA Safe Harbor guidelines. This dataset offers a unique resource for analyzing the clinical trajectories of patients with severe pneumonia.

## Background & Summary

Pneumonia is the largest cause of infectious deaths worldwide^1^ and one of the most common causes of hospital admissions in the United States.^2^ Patients who require ICU-level care are especially vulnerable. Community-acquired pneumonia (CAP), hospital-acquired pneumonia (HAP), and ventilator-associated pneumonia (VAP) are associated with high mortality, reported in some studies as high as 70%.^3^ The diagnosis and adjudication of pneumonia using structured data alone poses significant challenges due to the inherent complexity of the disease and limitations in how clinical information is documented and coded. Pneumonia is a clinical diagnosis that relies on a combination of symptoms (e.g., fever, cough, dyspnea), imaging findings (e.g., chest X-ray infiltrates), and laboratory results (e.g., leukocytosis or microbiological confirmation). However, none of these criteria are perfect, and each has limitations in sensitivity and specificity. For example, clinical notes may contain subjective or incomplete information, imaging reports might not always be definitive, and microbiological data can be absent or inconclusive, especially if patients were treated empirically. Additionally, the use of ICD codes to identify pneumonia cases may lead to misclassification, as coding practices often reflect billing priorities rather than clinical precision.^4^ Confounding factors such as coexisting conditions (e.g., heart failure or alveolar hemorrhage) that mimic pneumonia symptoms or have disease overlap further complicate adjudication. However, manual review by expert physicians takes significant effort, and significant heterogeneity exists on how physicians label pneumonia, even in clinical trials.^5^

Electronic health records (EHR) have revolutionized healthcare and are now present in most hospital systems. The intensive care unit (ICU) is an especially data-rich location, with numerous streams of data (continuous monitoring devices, ventilator parameters, etc.), of which the EHR is a key component.^6^ ICU EHR data typically include a wide array of information, such as vital signs, laboratory results, medication administration, clinical notes, and demographic details. These data facilitate a holistic view of a patient’s clinical trajectory, offering opportunities to improve understanding and promote better outcomes. Large ICU databases, such as MIMIC^7^, eICU^8^, and others,^9^ made available to researchers, have shown the vast array of knowledge that can be gleaned from these data. However, parsing these raw tables and modeling longitudinal data requires significant expertise, and as a result, many traditional approaches to analyzing ICU patient care often focus on features at the time of presentation and outcomes at discharge, collapsing the myriad events occurring during a patient’s stay. While useful, this approach has notable limitations. It tends to overlook the dynamic and evolving nature of critical illnesses, where the course of disease progression and intercurrent events, such as ICU complications or therapeutic interventions, play a significant role in patient outcomes. This is particularly problematic for patients with prolonged ICU stays, where key developments during their care trajectory may be poorly captured or entirely missed by admission-to-discharge predictive models.

To address this gap, the *CarpeDiem* framework was developed to examine clinical features on a day-to-day basis, mirroring the routine practice of daily multidisciplinary rounds.^10^ Daily rounds are a cornerstone of ICU care, during which the team reviews patient progress, examines vitals and laboratory results, and make major clinical decisions. Given that most laboratory tests are performed at least once daily and significant interventions often align with the daily clinical decision-making process, this time frame was chosen to organize and analyze patient data. By structuring data into daily bins, *CarpeDiem* provides a granular view of the patient journey, capturing the dynamic interplay of clinical features, complications, and interventions over time. This approach allows researchers to focus not only on initial diagnoses but also on the evolving landscape of patient care, enabling the study of intercurrent events that frequently define the outcomes of critically ill patients. We furthermore overlay a rich layer of expert clinician adjudication data for when pneumonia occurred, its category, and whether it was successfully or unsuccessfully treated. An example patient trajectory with different key timepoints is presented in **Figure 1**.

**Figure 1.**
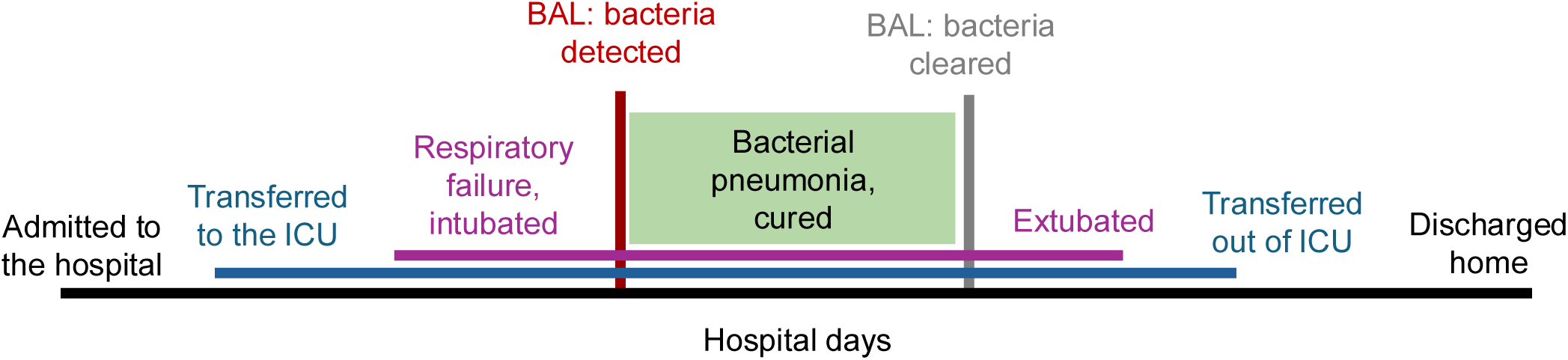
Example timeline of a patient’s hospital course. Key clinical events including hospital admission, ICU transfer, respiratory failure with intubation, extubation, ICU discharge, and hospital discharge are shown along a temporal axis. Bronchoalveolar lavage (BAL) sampling points are indicated, including initial detection of bacteria and subsequent clearance, with the interval of bacterial pneumonia highlighted and adjudication of ‘cured.’

Here, we describe the release of *CarpeDiem v1.9*, with additional features and patients compared with *CarpeDiem v1.0 and CarpeDiem v1.8*.^10,11^ We describe our pipeline for aggregating per-day clinical features, bronchoalveolar lavage (BAL) data, as well as episodes of pneumonia adjudicated using an independent, multi-reviewer process by an expert clinical team.^12^ The dataset provides granular, de-identified information compliant with Health Insurance Portability and Accountability Act (HIPAA) Safe Harbor rules, and is available to credentialed users via PhysioNet.^13^

## Methods

The data for this study were extracted from the EHR of patients enrolled in the Successful Clinical Response in Pneumonia Therapy (SCRIPT) study.^14^ SCRIPT is a prospective cohort study of patients with respiratory failure requiring mechanical ventilation, who undergo bronchoalveolar lavage (BAL) to diagnose and determine the etiology of known or suspected pneumonia. SCRIPT is approved by the Northwestern IRB (STU00204868). Patients’ families or licensed authorized representatives consent to take part in the study, which analyzes clinical data and BAL fluid to better characterize pneumonia pathophysiology and patient trajectories to improve future care. Additionally, the research team conducts interviews with patients or their families at enrollment and at the conclusion of the study, typically at hospital discharge. Patients are followed until the end of their hospitalization. Patients are given a new study identifier upon each hospitalization, so a small subset (<15) of patients is represented more than once in the dataset. Data were collected from patients enrolled between June 2018 and May 2023, which encompasses the entirety of patients enrolled in the ‘SCRIPT1’ cohort. Enrollment for its sequel study, SCRIPT2, or ‘SuperSCRIPT,’ remains ongoing as of March 2026.

### Clinical Cohort

All patients in our Northwestern Memorial Hospital (NMH) Medical Intensive Care Unit (MICU) who underwent BAL for the evaluation of known or suspected pneumonia, as indicated by abnormal chest imaging, abnormal vital signs or laboratory results, and clinician concern, were considered eligible for inclusion. These patients encompass a diverse range of underlying diagnoses, but all were severe enough to require mechanical ventilation. Exclusion criteria for the SCRIPT study include prior lung transplantation, prisoner status, pregnancy, and suspected pneumonia cases where bronchoscopy or nonbronchoscopic BAL is deemed unsafe by the attending physician. A small subset of patients underwent lung transplant during their hospitalization; these patients’ courses are censored at time of lung transplant and coded as having died.

### Data acquisition

The EHR data in this dataset is pulled from the Northwestern Medicine Enterprise Data Warehouse (EDW), which serves as the primary repository for clinical data at Northwestern Medicine.^15^ This system integrates approximately 150 distinct data sources, including the Epic EHR system, which are updated nightly. The data loading process uses Microsoft technologies such as Visual Studio and SQL Server Integration Services (SSIS) and is managed through scheduling via the SQL Server Job Agent. Following data integration, data engineers and architects within the EDW team consolidate these sources into datamarts using custom SQL scripts. Subsequently, analysts on the EDW team leverage these datamarts to develop validated reports, dashboards, and data extracts in collaboration with clinical experts. Extensive chart review was performed for each report to ensure validity of its output. Data spans basic EHR features such as vital signs, to more specialist features such as ventilator parameters, to more detailed composite scores such as the Narrow Antibiotic Therapy score.^16^ Clinical notes were extracted to examine immunocompromised features using a large language model pipeline, described further in our other work.^17^

The SCRIPT research team interviews patient families and reviews their medical record to gather additional information such as treatment goals, whether the patient is immunocompromised, and outcomes at the end of hospitalization that are difficult to abstract automatically from the EHR, such as changes in goals of care. Features derived from the research team are highlighted in the Data Dictionary. Data collected by the research team are manually entered into our local secure REDCap instance.

### Clinical adjudication

A team of five Pulmonary and Critical Care attending physicians reviews each patient chart and adjudicates episodes of pneumonia using a predefined series of questions, with predefined guidelines. A chart is given to two independent reviewers, with discrepancies sent to a third independent reviewer. Additional discrepancies are reviewed by the entire team at a weekly meeting. These data include determining the type of pneumonia (or lack of pneumonia – ‘Non-Pneumonia Control/NPC’), the duration of the pneumonia episode for bacterial pneumonia, and whether the pneumonia was successfully cured, not cured, or indeterminate. Pneumonia episode information is aligned relative to the BAL whose results define that episode. Additional details of the review process, including agreement and adjudication questions, are available in our previous work.^12^

### Data transformation and joining

All relevant data in the dataset are presented in a single table for convenience. The primary dimension of this table is patient hospital days, and the values for each hospital day for each patient are derived from the data sources described above according to the following logic.

Each patient-hospital-day is a row in the dataset, and pneumonia episode data and BAL data are joined to it. Demographic and outcome data are repeated for each hospital-day for the same patient. EHR data is aggregated for multiple measurements per day via mean and minimum or maximum, with the directionality depending on which direction is worse. Outlier handling is performed before aggregation using a predetermined set of physiologically reasonable ranges. Textual results are removed, and only numerical values are used for analysis. Greater than or less than signs are removed, and the upper/lower bounds are used as numerical values. Units are harmonized after examination for distribution and conversions as needed (for example, converting lactate from mg/dL to mmol/L by dividing by 9).

Pneumonia episode data are recorded for the patient-hospital-day when the episode-defining BAL was performed. The physician adjudication team evaluates the patient’s status on days 7-8, 10, and 14 days after the episode-defining BAL. Cured cases have durations of 7, 10, or 14 days based on the corresponding recorded evaluation, or shorter based on patient extubation after the episode-defining BAL. Indeterminate cases use the last available reading (7, 10, or 14 days) or the day antibiotics stopped. Persistent or superinfected cases are not cured, with durations based on the latest occurrence (7, 10, or 14 days), when antibiotics stopped or hospital discharge. Non-pneumonia controls are labeled separately, and remaining cases are classified as final episodes with success status and duration extending to the last day. Viral-only episodes are excluded from the evaluation of clinical cure, with durations reset to NA due to lack of clear definition of viral cure. Additional detailed logic is developed using a series of rules that are documented in detail in our code repository, available on GitHub.

Data is synthesized from EDW data that comes from several core reports that are refreshed monthly. On top of this, we have built out several custom reports for specific projects. The research team enters data via REDCap, and reports are exported from that monthly. Issue tracking is managed using GitHub and Smartsheet, supplemented by manual checks and fixes based on the front-facing EHR, EPIC. Any data-related questions encountered by researchers during analysis are logged as new issues in GitHub. Additionally, new parameters requiring inclusion in reports are logged in GitHub, assigned to data analysts or architects, and tagged with relevant labels (e.g., missing data, data error, documentation, or future needs) for priority management. A monthly data tracker is run to assess data quality, with a focus on cross-referencing subject IDs and sample IDs between the REDCap and clinical demographics forms. A heatmap is generated to visualize missing data, and detailed IDs are flagged for any mismatches between the clinical data and REDCap forms. These discrepancies are then reviewed, corrected, and documented in a Smartsheet tracking sheet, which includes the original value, the corrected value, and the reason for the update. Metadata are also systematically recorded and managed in Smartsheet, ensuring organized and accessible documentation for the SCRIPT team. The curated data are harmonized and then undergo deidentification checks before being published. Some internal projects require the use of a limited data set, including data such as dates and internal identifiers; an internal version of files with additional security controls are kept for this reason. **Figure 2** gives a broad overview of the data process.

**Figure 2.**
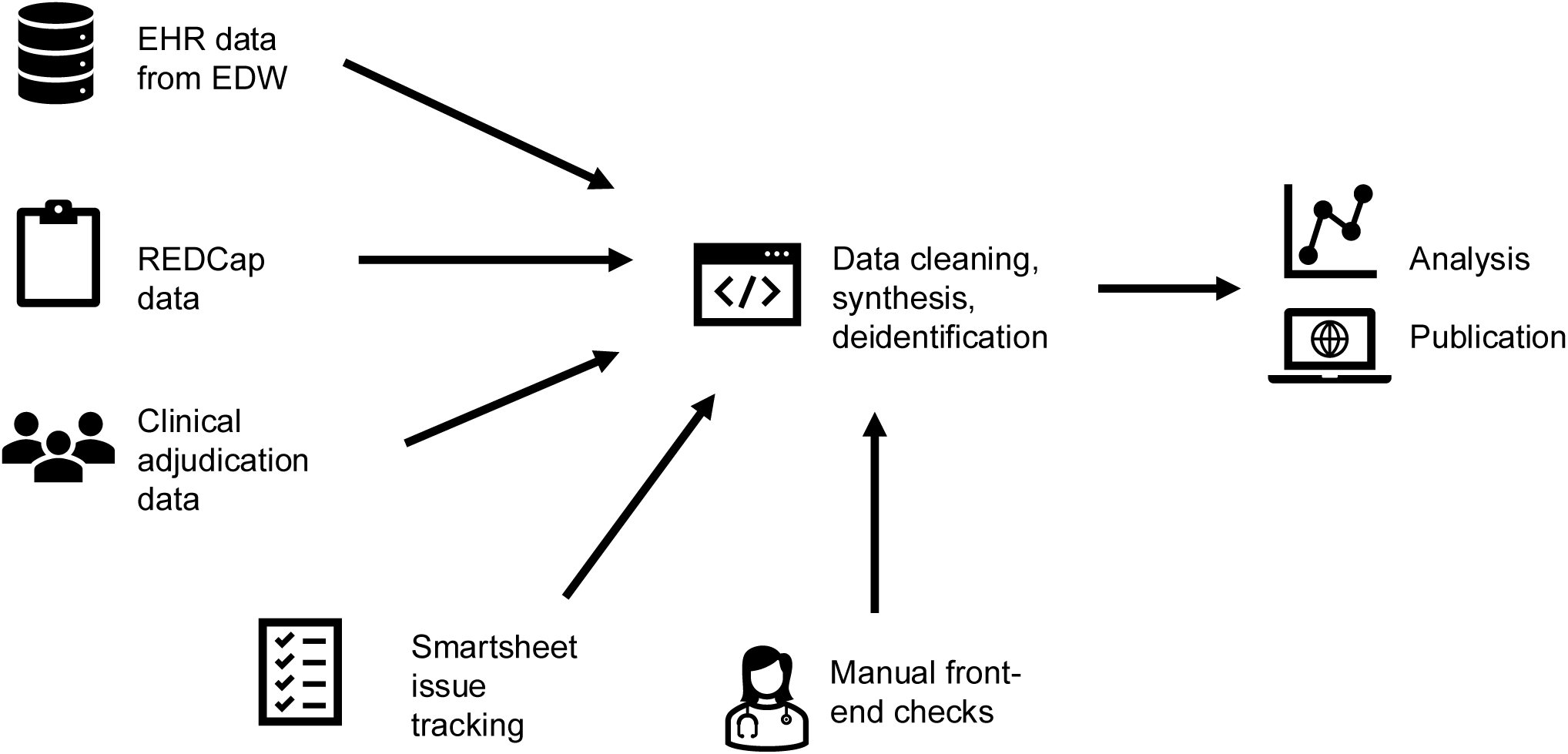
Overview of the data workflow. Multiple data sources including electronic health record (EHR) data from the enterprise data warehouse (EDW), REDCap surveys, clinical adjudication records, Smartsheet issue tracking, and manual front-end quality checks are integrated into a centralized pipeline for data cleaning, synthesis, and deidentification. The processed dataset is then used for downstream analysis and publication.

### Deidentification Protocol

To ensure compliance with privacy standards, the dataset was de-identified in accordance with the Health Insurance Portability and Accountability Act (HIPAA) Safe Harbor rules.^18^ All dates were removed and replaced with relative time points based on each patient’s hospital stay. For patients aged over 89, age data were aggregated into a single category labeled “91,” consistent with existing deidentification practices used by other databases such as MIMIC.^7^ Following de-identification by CAG, WTL, and NSM, two other authors (LVR, MK) independently reviewed the dataset to confirm compliance with Safe Harbor rules. This involved manual inspection of distinct values in the dataset, and analysis using the ARX data anonymization tool.^19^ Patients of a rare ethnicity with fewer than five were recoded to protect patient privacy.

To ensure no direct or indirect identifiers are accidentally left in the code repository, LVR developed a review protocol that leveraged three open-source tools, Presidio,^20^ Philter,^21^ and ClinDeID^22^ to flag parts of the source code for manual review. Human inspection is needed as names and dates associated with the source code authors are typically flagged as identifiers. Full paths and dates of file names were also redacted for security purposes. Work describing our deidentification protocol in greater detail is under development.

### Data Records

Access to the SCRIPT *CarpeDiem* Dataset v1.8 is provided to credentialed users who sign a data use agreement (DUA) via PhysioNet at: https://physionet.org/content/script-carpediem-dataset/1.8.0/. ^11^ Version 1.9 has been submitted and is under review. This dataset is organized at the level of a single patient-hospital-day, with each row capturing one day of a patient’s hospitalization and enriched with admission summary information. The first 44 columns contain demographic characteristics and overall outcomes summarized across the patient’s entire hospital stay. The next 113 columns record day-by-day clinical measurements, providing a detailed view of how key patient features evolve over time. The final 39 columns focus on events specific to that hospital day, including bronchoalveolar lavage data when performed and adjudication of pneumonia episodes, such as episode type and associated outcomes.

### Demographics

Information includes age at admission, sex, race, ethnicity, body mass index (BMI), smoking status, admission source, and comorbidities including immunocompromised status. There are 704 patients in the cohort with median [IQR] age of 62 [51,71]. 60% of patients were male, 20% Black or African American, and 20% Hispanic/Latino. The median [IQR] intubated days was 11 [5,25]; 45% of patients had an unfavorable outcome (death, discharge to hospice, or requiring lung transplantation). **Figure 3** summarizes basic demographics and outcomes for the cohort.

**Figure 3.**
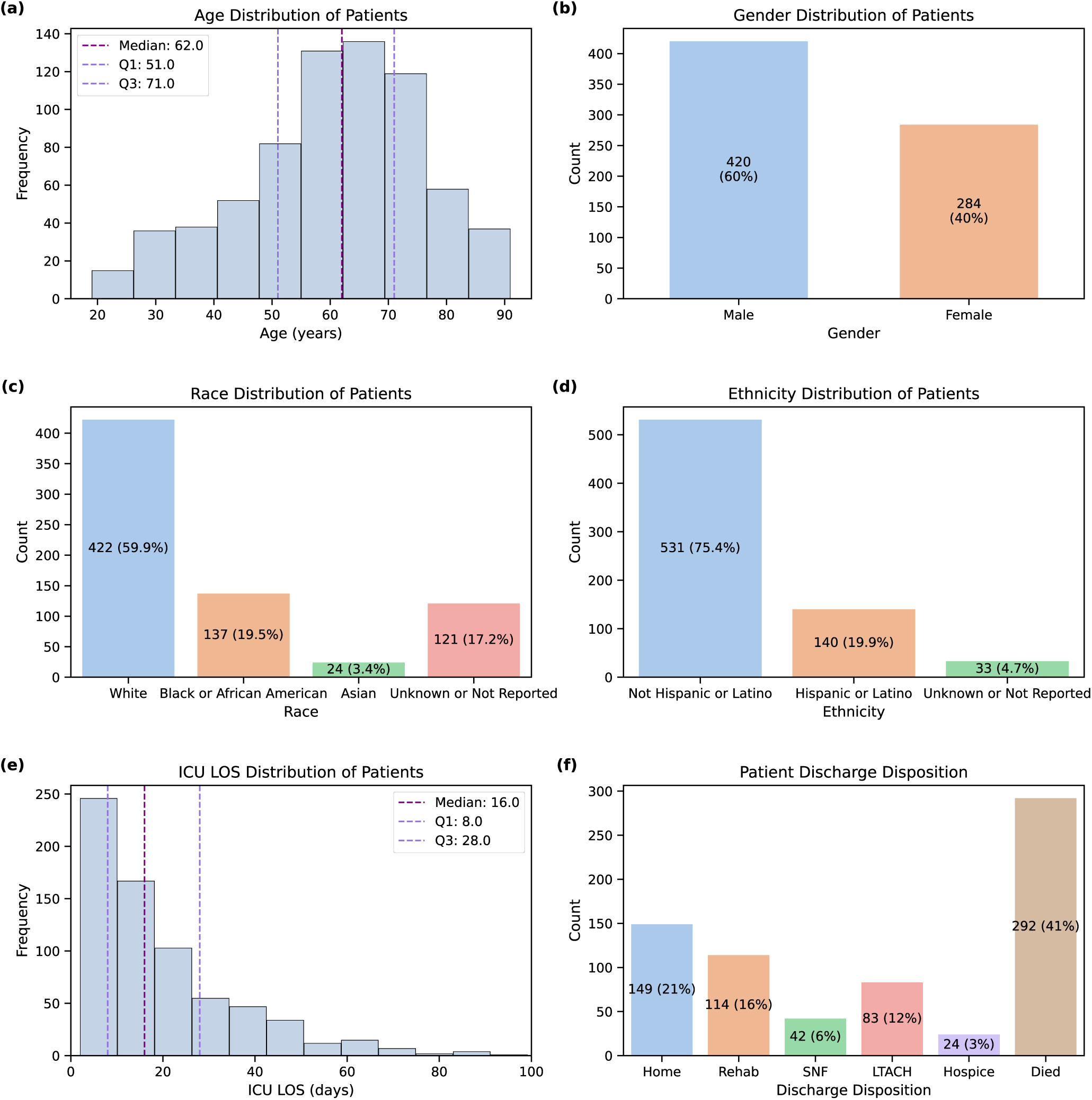
Demographics and clinical outcomes of the study cohort. Panels show distributions of (a) age, with median and interquartile range indicated; (b) sex; (c) race; (d) ethnicity; (e) intensive care unit (ICU) length of stay (LOS); and (f) discharge disposition. Counts and percentages are annotated where applicable, summarizing baseline characteristics and hospital outcomes for the cohort.

### Daily features

These span clinical features aggregated on a per-day level via mean and either min/max, see Data Dictionary for full details. We selected clinical features considered representative of those typically evaluated by physicians during daily multidisciplinary ICU rounds. These features encompassed:

- **Vital signs**: temperature, heart rate, systolic blood pressure, diastolic blood pressure, mean arterial pressure, respiratory rate, and oxygen saturation;
- **Hemodynamic parameters**: norepinephrine dose (mcg/kg/min), lactic acid;
- Support devices: extracorporeal membrane oxygenation (ECMO), acute renal replacement therapy (hemodialysis [HD], continuous renal replacement therapy [CRRT]);
- **Mental status evaluation**: Glasgow Coma Scale (GCS) subscores (eye opening, motor response, verbal response), and Richmond Agitation Sedation Scale (RASS);
- **Respiratory support**: status of intubation, positive end-expiratory pressure (PEEP), FiO2, plateau pressure, minute ventilation, driving pressure;
- **ABG**: pH, P_a_O_2_, P_a_CO_2_, P_a_O_2_/F_i_O_2_ ratio;
- **Labs**: White blood cell count (WBC), including lymphocytes, neutrophils, and eosinophils, hemoglobin, platelets, red cell distribution width (RDW), sodium, bicarbonate, blood urea nitrogen (BUN), creatinine, glucose, albumin, bilirubin, prothrombin time (PT), partial thromboplastin time (PTT), C-reactive protein (CRP), D-dimer, ferritin, lactate dehydrogenase (LDH), and procalcitonin;
- **Medication**: narrow antibiotic therapy (NAT) score, hydrocortisone-equivalent steroid dose, whether patient received tocilizumab, sarilumab, remdesivir, or a study drug; cumulative dose of steroids and cumulative NAT score.

### Bronchoalveolar lavage (BAL) features

- Whether the patient had a BAL this date;
- BAL cell count differential for: neutrophils, macrophages, monocytes, lymphocytes, other cells;
- Pathogen results: whether a virus, bacteria, fungi, or resistance marker was recovered; full pathogen data is formatted in JSON as detailed in our code repository.

### Pneumonia episode adjudication

- Each episode is classified by pneumonia category (community, hospital, ventilator-associated, or non-pneumonia control) and by etiology (bacterial, viral, mixed, microbiology-negative, or other);
- Adjudicators determine whether the episode was cured, document the evidence used, and record reasons for treatment failure when applicable;
- Episode length is calculated for bacterial and mixed infections, with clinician assessments of episode status at standardized follow-up time points;
- For episodes judged not to represent pneumonia, alternative causes of pulmonary infiltrates, fever, leukocytosis, or other infections are documented.

### Adjudication of pneumonia episodes

A panel of critical care physicians adjudicated pneumonia cases using bronchoalveolar lavage results as the primary diagnostic evidence. Based on this review, patients were classified into four groups: non-pneumonia controls, COVID-19, other viral pneumonia, and other pneumonia. In addition, the panel identified discrete pneumonia episodes for each patient and assigned an outcome to each episode, categorizing it as successfully cured, indeterminate, or not cured. Detailed descriptions of the adjudication process are provided in related manuscripts.^12^

210 patients had COVID-19, 67 had pneumonia secondary to other respiratory viruses, 317 had other pneumonia (bacterial), and 114 were initially suspected of having pneumonia yet subsequently adjudicated as having respiratory failure unrelated to pneumonia (non-pneumonia controls). There are 153 episodes of community-acquired pneumonia (CAP), 265 episodes of hospital-acquired pneumonia (HAP), and 417 episodes of ventilator-associated pneumonia (VAP). Patients had from one to eight episodes, and a Sankey plot demonstrating episode flow is presented in **Figure 4a** for type of pneumonia, and in **Figure 4b** for pneumonia etiology. 390 episodes were adjudicated to be successfully cured, 168 were adjudicated as indeterminate, and 277 were adjudicated as not cured. 44.89% of the cohort had an unfavorable outcome defined as discharge to hospice or death.

**Figure 4.**
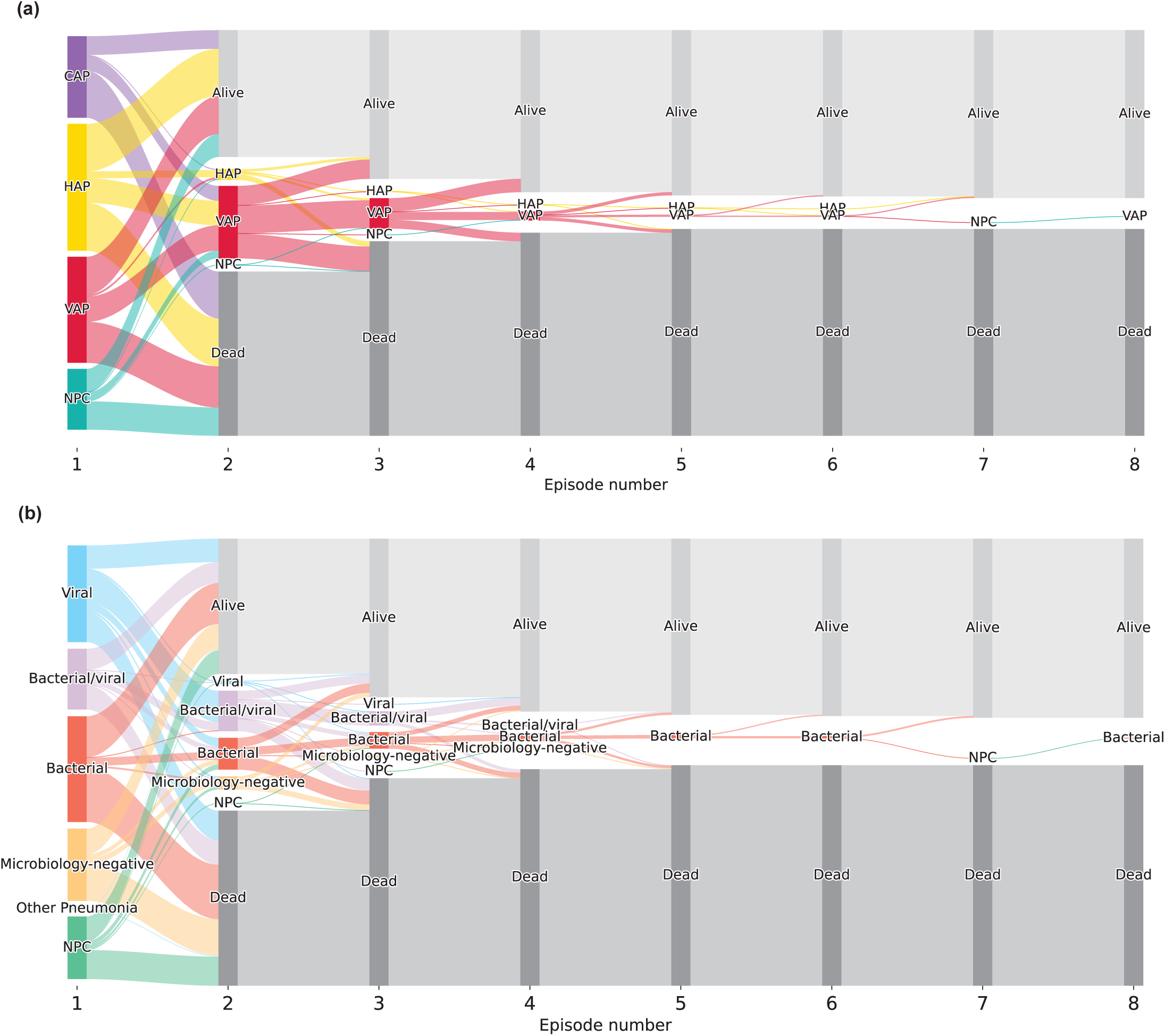
Sankey diagrams illustrating transitions in pneumonia episodes over time. Panel (a) depicts patient flow across successive episodes by pneumonia type (community-acquired, hospital-acquired, and ventilator-associated pneumonia). Alive and dead represent patient discharge. Panel (b) shows transitions by pneumonia etiology (viral, bacterial, mixed bacterial–viral, microbiology-negative, non-pneumonia conditions, and other pneumonia). Vertical bands represent episode number, with flows indicating progression between categories and clinical outcomes (discharged alive or deceased).

Clinical and outcome data are included for each patient, capturing measures, such as discharge disposition, cumulative days in the ICU, cumulative days of intubation, the number of ICU stays, and whether a tracheostomy was required. Discharge disposition is classified into six categories, in order of favorability: home, acute inpatient rehabilitation, skilled nursing facility, long-term acute care hospital, hospice, or death. Patients who underwent lung transplantation during their ICU stay were classified as having died, reflecting replacement of the native lungs and the high likelihood of mortality in the absence of transplantation.

### Technical Validation

The SCRIPT *CarpeDiem* database was built upon several years of multidisciplinary collaboration between clinicians and scientists across several core groups within the SCRIPT Systems Biology Center (Modeling, Database Management, Administrative, Clinical). All groups held regular meetings, in addition to monthly meetings with the entire group. Issues and projects were tracked using a standardized system, code was managed through Git repositories to ensure reproducibility.

Numerous manual integrity checks were performed by clinicians on the front end EHR to ensure data reports were accurate. Each data source for this database (EDW, REDCap, adjudication data) has their own independent manual validation systems where data issues are recorded and addressed.

The distribution of each feature in the dataset was manually inspected for the outliers and the corresponding data frequency and missingness patterns. Additional checks were run to check the regular measurement of features such as vitals, compared with less common features such as WBC, and less common occurrences such as BALs. As we worked with the data, we discovered and fixed numerous bugs and edge cases. Greater than 50 patients were reviewed for consistency between our processed dataset and the hospital EHR system to ensure quality. Deidentification checks of both the data, and the code repositories, were performed, see above Methods for additional details.

### Usage Notes

This dataset has been used to explore daily clinical states through unsupervised clustering of patient-day features. Using this approach, 14 distinct clusters with unique clinical characteristics were identified, providing a granular map of patient trajectories.^10^ This clustering framework was employed to investigate patterns in pneumonia progression and outcomes, comparing patients with and without COVID-19. The code repository used in these analyses is publicly available on GitHub,^23^ and users are encouraged to raise issues or questions on that platform. We have furthermore leveraged this dataset to examine the antibiotic de-escalation patterns across different types of pneumonia,^24,25^ the characteristics of BAL % neutrophils in patients with neutropenia and immunocompromise,^26,27^ predict next-day extubation,^28,29^ and examine different BAL galactomannan cutoffs in patients who grew *Aspergillus*.^30,31^

A note on missing data: as with many datasets derived from electronic health records (EHRs), missing data are a notable limitation. Nevertheless, these gaps may be informative, reflecting clinical decision-making processes. For example: physicians may choose not to perform certain laboratory tests if they are deemed clinically unnecessary or uninformative; missing data on ventilator parameters indicate that the patient has been extubated, reflecting the absence of a ventilatory support device rather than a data collection error. See **Figure 5** for data presence/absence across the dataset.

**Figure 5.**
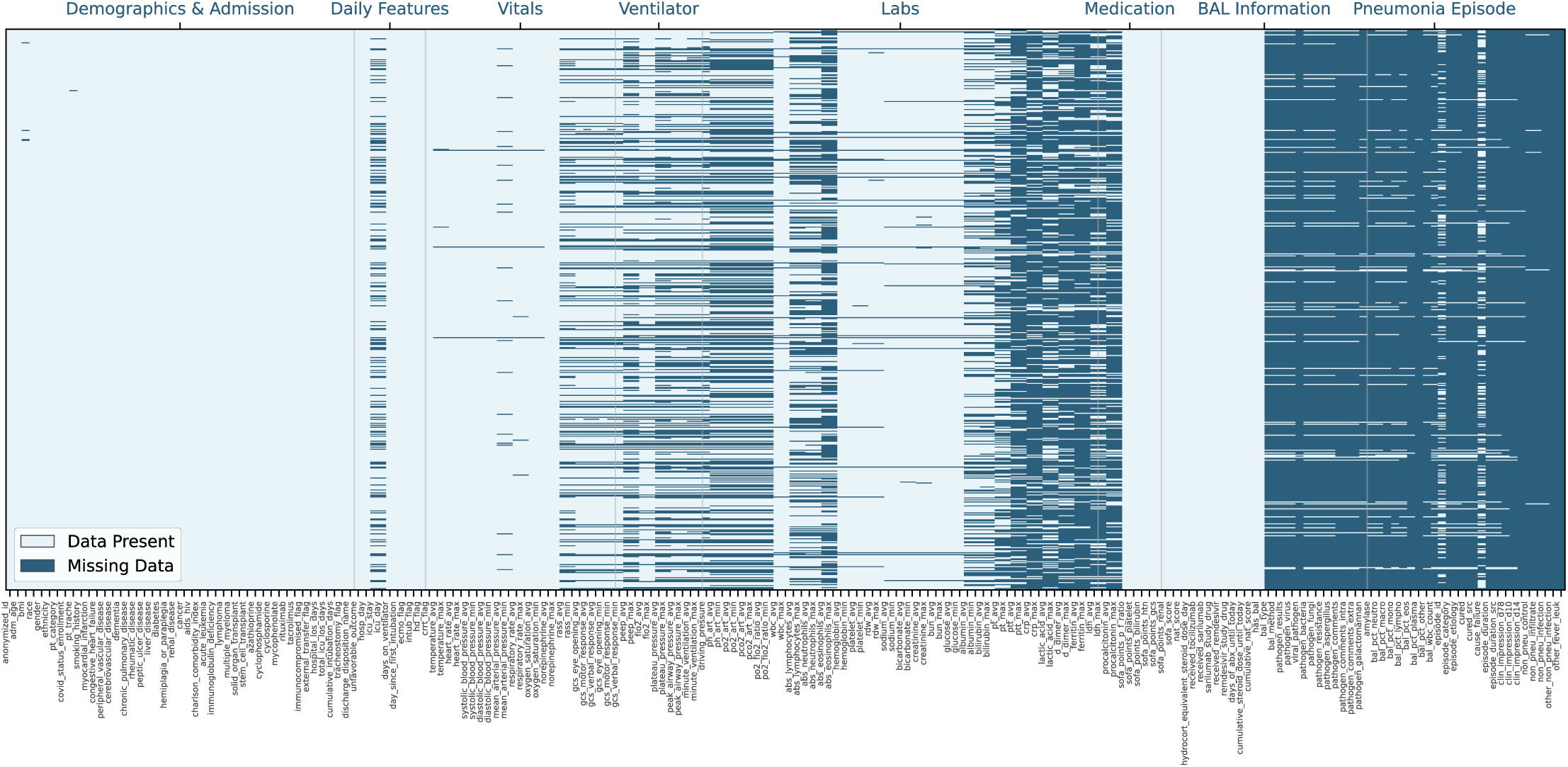
Data completeness across clinical domains. Heatmap showing presence and absence of variables across demographics and admission characteristics, daily features, vital signs, ventilator settings, laboratory values, medications, BAL information, and pneumonia episode labels. Missingness reflects clinical context rather than random loss, for example ventilator data are absent after extubation and BAL results are present only on days when a BAL was performed.

Project specific code repositories can be found within our organization repository.^32^ This dataset opens many potential avenues for innovative studies, such as exploring associations between clinical states and treatment responses, developing dynamic predictive models to understand how daily changes in patient conditions influence long-term outcomes, and uncovering latent trends through unsupervised learning and clustering techniques. Researchers are encouraged to leverage the provided tools, including the GitHub repository,^32^ to conduct these and other analyses. The detailed documentation and accompanying browser further simplify data exploration and analysis, supporting a wide range of clinical and research applications. Users are encouraged to raise questions and track issues via GitHub as well.

## Supporting information

NU SCRIPT Study Investigators

## Data Availability

The majority of SCRIPT data has already been made available to credentialed users who sign a DUA via PhysioNet: https://physionet.org/content/script-carpediem-dataset/1.8.0/

Measurement(s)

Electronic Health Record, clinical

Technology Type(s)

Electronic Health Record

Sample Characteristic - Organism

*Homo sapiens*

Sample Characteristic – Environment

Hospital

Sample Characteristic – Location

Chicago, Illinois, USA

## Code Availability

The code to build this dataset is available on GitHub: https://github.com/NUSCRIPT/carpediem_v1.9/. Example usage of the dataset in projects are also available in project repositories such as https://github.com/NUSCRIPT/CarpeDiem, https://github.com/NUSCRIPT/gao_aspergillus_2024, https://github.com/NUSCRIPT/mz_abx_deescalation_2024, amongst others in our organization repository.

## Acknowledgements

LLMs were used to help coding, drafting, and editing for grammar/clarity, with all output reviewed by authors; authors take responsibility for final output.

The SCRIPT Study is funded by NIH NIAID U19AI135964.

Work in the Division of Pulmonary and Critical Care is also supported by NUCATS, SQLIFTS, and the Canning Thoracic Institute of Northwestern Medicine.

CAG is supported by NIH/NHLBI K23HL169815, a Parker B. Francis Opportunity Award, and an ATS Unrestricted Grant.

NSM is supported by AHA 24PRE1196998 (https://doi.org/10.58275/AHA.24PRE1196998.pc.gr.190609).

CIP is supported by NIAID (U19AI135964), Northwestern University Clinical and Translational Sciences Institute (5KL2TR001424-09).

G.R.S.B. was supported by a Chicago Biomedical Consortium grant, Northwestern University Dixon Translational Science Award, Simpson Querrey Lung Institute for Translational Science, the NIH (grant nos. P01AG049665, P01HL154998, U54AG079754, R01HL147575, R01HL158139, R01HL147290, R21AG075423 and U19AI135964) and the Veterans Administration (award no. I01CX001777).

A.V.M. was supported by the NIH (grant nos. U19AI135964, P01AG049665, P01HL154998, U19AI181102, R01HL153312, R01HL158139, R01ES034350 and R21AG075423).

BDS is supported by the NIH (R01HL149883, R01HL153122, P01HL154998, P01AG049665, and U19AI135964).

RGW is supported by NIH grants (U19AI135964, U01TR003528, P01HL154998, R01HL14988, and R01LM013337).

The funding sources did not have a role in the design, execution, or prior review of the study or in the data presented in this manuscript. Opinions expressed in this work do not necessarily reflect those of the funding sources.

## Author contributions

CAG performed programming and analysis, including data cleaning, data stitching, visualizations, repository maintenance, issue management, participated in clinical pneumonia episode adjudication, performed significant chart reviews for unusual cases, and drafted the manuscript. NSM performed programming and analysis, data cleaning, data stitching, visualizations, repository maintenance, and issue management. MK compiled raw data from the EDW and certified deidentification while also assisting with data management and issue management. AP compiled raw data from the EDW. LVR certified deidentification and was involved in data management, data curation, and issue management. WTL performed programming and analysis, data cleaning, data stitching, repository maintenance, and issue management. PN compiled raw data from the EDW. VG participated in significant chart reviews for unusual cases and adjudicated immunocompromised status from clinical notes using an LLM pipeline. HKD oversaw the research team that enrolled patients and gathered clinical data. RKC designed the adjudication entry system and compiled clinical adjudication data. CP participated in clinical pneumonia episode adjudication. NN participated in clinical pneumonia episode adjudication. DS assisted with data management. TS assisted with data management. JS assisted with data management and provided guidance and supervision. TW assisted with data management and provided guidance and supervision. RGW participated in clinical pneumonia episode adjudication, obtained funding, and provided guidance and supervision. GRSB was involved in obtaining funding and provided guidance and supervision. AVM helped secure funding and contributed to guidance and supervision. BDS participated in clinical pneumonia episode adjudication, obtained funding, as well as providing guidance and supervision. All authors read and approved the final manuscript.

